# Genomic determinants of response and resistance to inotuzumab ozogamicin in B-cell ALL

**DOI:** 10.1101/2023.12.06.23299616

**Authors:** Yaqi Zhao, Nicholas J Short, Hagop M Kantarjian, Ti-Cheng Chang, Pankaj S Ghate, Chunxu Qu, Walid Macaron, Nitin Jain, Beenu Thakral, Aaron H Phillips, Joseph Khoury, Guillermo Garcia-Manero, Wenchao Zhang, Yiping Fan, Hui Yang, Rebecca S Garris, Lewis F Nasr, Richard W Kriwacki, Kathryn G Roberts, Marina Konopleva, Elias J Jabbour, Charles G Mullighan

**Affiliations:** Department of Pathology, St. Jude Children’s Research Hospital, Memphis, TN; Center of Excellence for Leukemia Studies, St. Jude Children’s Research Hospital, Memphis, TN; Department of Leukemia, MD Anderson Cancer Center, Houston, TX; Center for Applied Bioinformatics, St. Jude Children’s Research Hospital, Memphis, TN; Department of Hematopathology, MD Anderson Cancer Center, Houston, TX; Department of Structural Biology, St. Jude Children’s Research Hospital, Memphis, TN; Department of Pathology and Microbiology, University of Nebraska Medical Center, Omaha, NE; Department of Medicine/Oncology and Molecular Pharmacology, Albert Einstein College of Medicine, Bronx, NY

## Abstract

Inotuzumab ozogamicin (InO) is an antibody-drug conjugate that delivers calicheamicin to CD22-expressing cells. In a retrospective cohort of InO treated patients with B-cell acute lymphoblastic leukemia, we sought to understand the genomic determinants of response to InO. Acquired *CD22* mutations were observed in 11% (3/27) of post-InO relapsed tumor samples. There were multiple *CD22* mutations per sample and the mechanisms of CD22 escape included protein truncation, protein destabilization, and epitope alteration. Hypermutation by error-prone DNA damage repair (alternative end-joining, mismatch repair deficiency) drove CD22 escape. Acquired loss-of-function mutations in *TP53*, *ATM* and *CDKN2A* were observed, suggesting compromise of the G1/S DNA damage checkpoint as a mechanism of evading InO-induced apoptosis. In conclusion, genetic alterations modulating CD22 expression and DNA damage response influence InO efficacy. The escape strategies within and beyond antigen loss to CD22-targeted therapy elucidated in this study provide insights into improving therapeutic approaches and overcoming resistance.

**KEY POINTS:** We identified multiple mechanisms of CD22 antigen escape from inotuzumab ozogamicin, including protein truncation, protein destabilization, and epitope alteration.

Hypermutation caused by error-prone DNA damage repair was a driver of CD22 mutation and escape.

**VISUAL ABSTRACT:** 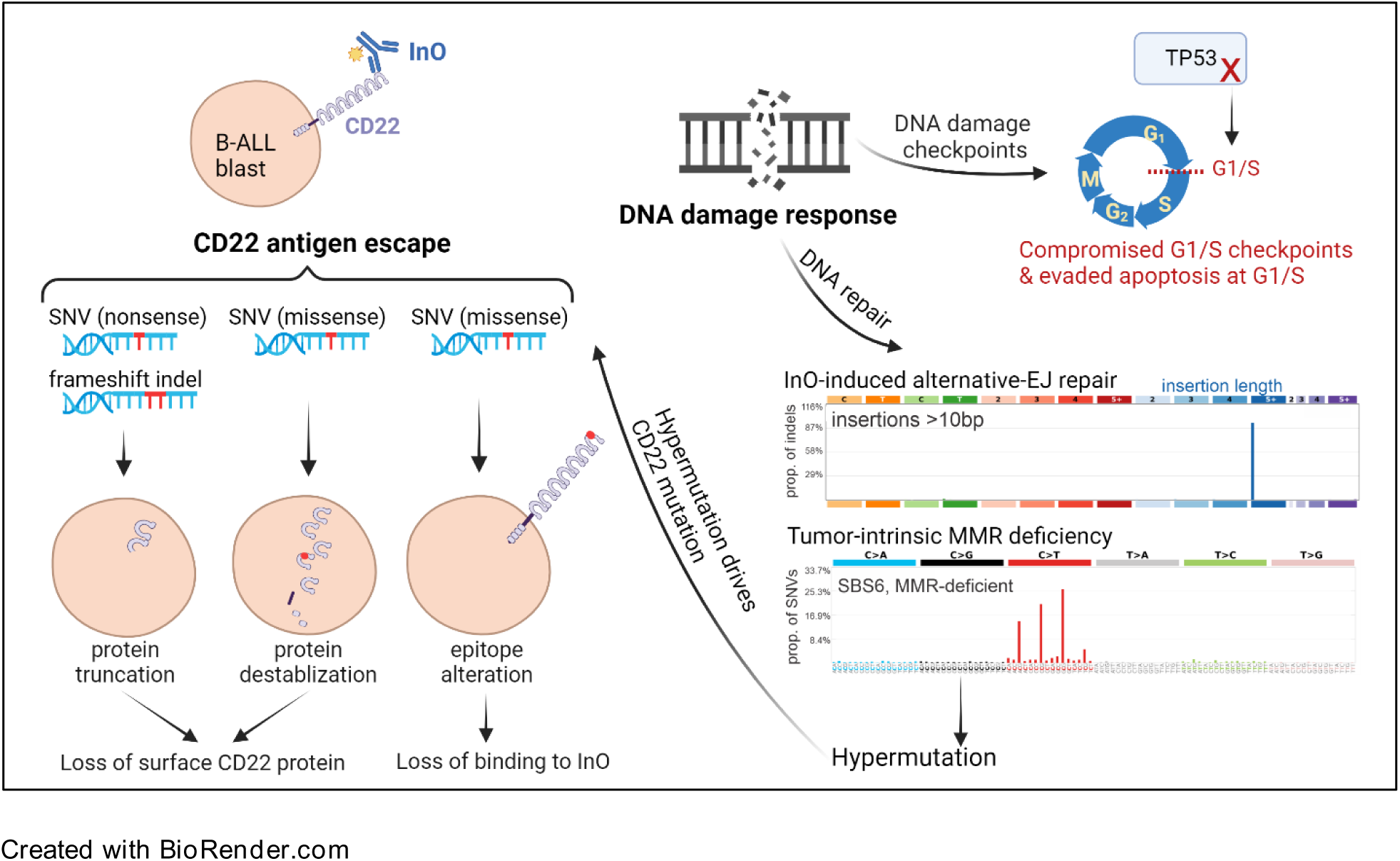

## INTRODUCTION

Inotuzumab ozogamicin (InO) is a highly active antibody-drug conjugate for the treatment of B-cell acute lymphoblastic leukemia (ALL). InO was designed to deliver the chemotherapeutic payload calicheamicin to cells expressing CD22, atype I transmembrane protein solely expressed on B-lineage cells.^1,2^ Calicheamicin is a highly potent toxin that binds to the DNA minor grove and induces double-stranded DNA breaks (DSBs).^3^ InO elicits high rates of remission and undetectable measurable residual disease (MRD), and represents an advance in therapeutic options for relapsed/refractory (R/R) B-cell ALL.^4–6^ Despite the established survival advantage of InO compared with conventional chemotherapy, some patients are refractory to InO; even among responding patients, InO monotherapy is rarely curative. Elucidating the biological and genomic determinants of response is therefore imperative to develop rationale strategies to overcome this resistance.

Delivery of the calicheamicin payload requires CD22 binding and internalization. Loss or downregulation of target antigen is one way by which tumor cells escape targeted immunotherapy. For example, truncating mutations and alternative splicing in *CD19* were shown to lead to antigen loss following therapy with CD19-directed chimeric antigen receptor (CAR)-T cell and blinatumomab.^7–9^ Dysregulated *CD22* splicing was reported to be a mechanism of epitope downregulation and resistance to CD22-directed immunotherapies.^10^ However, other potential mechanisms of CD22 antigen escape and therapy resistance are poorly understood.

Upon the successful delivery of calicheamicin to target cells, cytotoxicity of InO relies on its ability to produce DSBs, and this DNA damage can activate multiple apoptotic mechanisms.^5^ The known pathways of DSBs repair include the high-fidelity homologous recombination (HR), and error-prone repairs such as non-homologous end joining (NHEJ), alternative end-joining (a-EJ) and single-strand annealing (SSA).^11^ When repairing non-blunt ends DSBs like InO-induced damage,^12^ the error-prone repairs result in distinct types of genomic sequence “scar”: NHEJ creates small insertions and deletions; a-EJ creates longer (>10 nucleotides) insertions; SSA is prone to generate deletions and translocations.^11^

There are multiple DNA damage checkpoints during the cell cycle: G1/S checkpoint, G2/M checkpoint, and a less well-defined intra-S checkpoint.^13^ Of these checkpoints, G1/S checkpoint is unique in depending primarily on the function of p53 .^13,14^ Previous study in cell lines reported cells surviving InO were mostly blocked in the G2/M phase, suggesting the reliance on G2/M checkpoint in response to InO-induced DNA damage.^15^

In this study, we performed a comprehensive investigation of mechanisms of escape from InO therapy. We interrogated patients’ genomic data to examine CD22 escape strategies, how tumor cells overcome InO-induced DSBs, and the usage of DNA damage checkpoints during InO resistance; we also utilized genome wide CRISPR screens *in vitro* to identify novel targets of InO resistance.

## RESULTS

### Patient characteristics and response to InO by subtype

We studied 85 adult patients with R/R (n=81, 95%) or newly diagnosed (n=4, 4.7%) B-ALL treated with InO monotherapy (n=55, 65%) or InO in combination with low-intensity chemotherapy (n=30, 35%). This was a retrospective study, and the inclusion criterion was availability of pre-InO and/or post-InO tumor samples, either as banked cells or nucleic acids from bone marrow or peripheral blood-derived tumor cells. All patients had detectable disease (>5% blasts) in the bone marrow at the time of receiving treatment. Demographic and clinical features are summarized in Table 1. The median age was 42 years (range 18-84). The median number of prior therapies was two (range 0-5) and 55% patients received InO as their second or subsequent course of salvage therapy. The cohort was divided into responders (those who achieved complete remission [CR, n=26] or complete remission with incomplete hematologic recovery [CRi, n=25]) and non-responders (NR, either no response [n=31] or early death [n=3]). Thirty-one (61%) responders subsequently underwent allogeneic hematopoietic stem cell transplantation (HSCT) after attaining remission with InO (Figure 1, Supplementary Table 1). Of the 35 patients with post-InO relapse, 11 (31%) relapsed during InO therapy, 7 (20%) after completion of InO without undergoing HSCT, and 17 (49%) after completion InO and after subsequent HSCT. Samples were obtained pre-InO only (n=42), pre-& post-InO (n=28), and post-InO only (n=15). The post-InO samples included samples from patients who achieved CR/CRi but subsequently relapsed (n=27), and patients who were refractory to InO (n=16).

**Figure 1.**
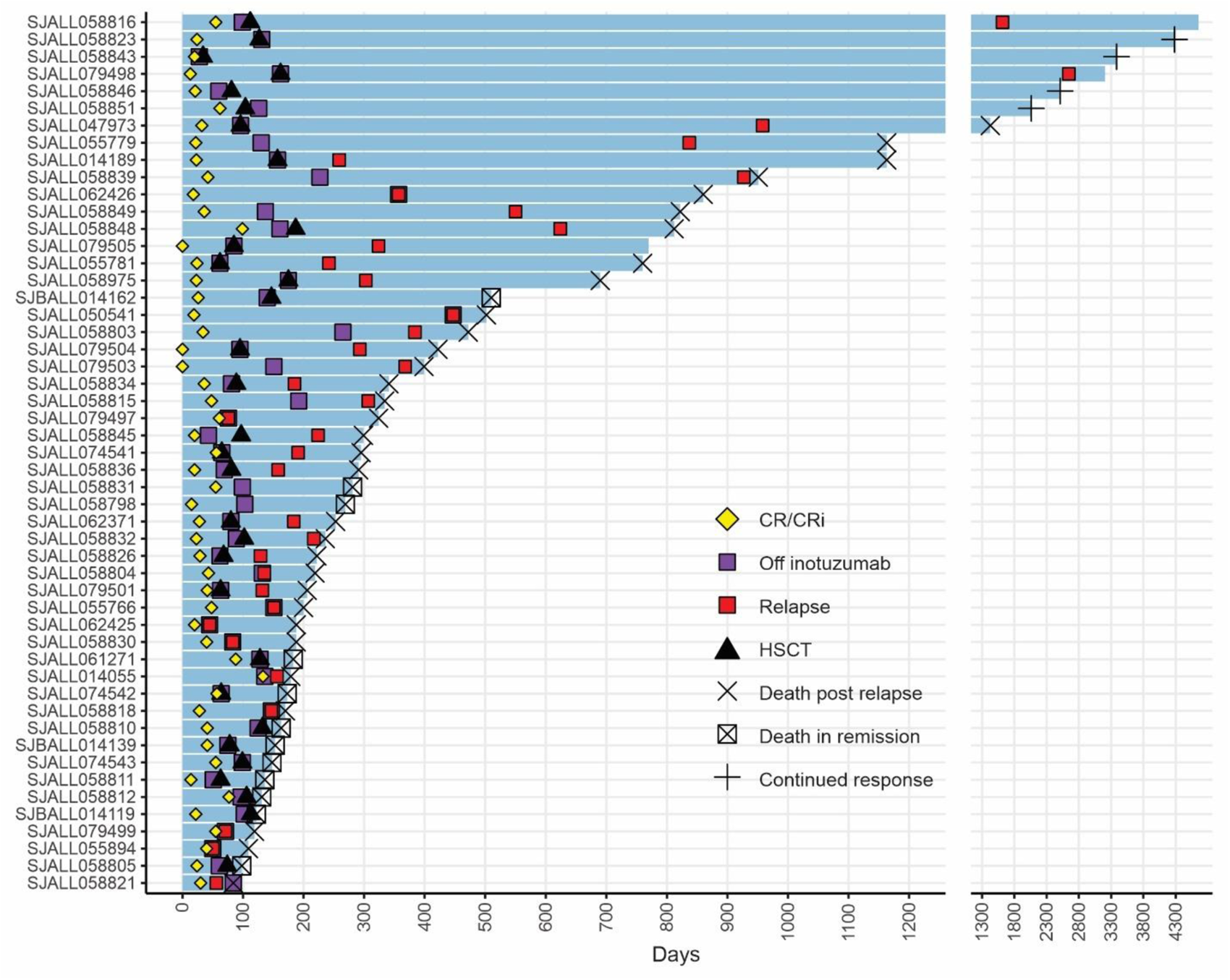
Response to inotuzumab. Swimmer plot for InO responders (n=51). Each bar represents from the start of InO therapy to last follow up. Once attaining CR/CRi, patients were off InO therapy, and 31 responders subsequently received hematopoietic stem cell transplantation (HSCT).

**Table 1.** Patient demographics and clinical characteristics (N = 85)

|  | **n (%)** |
| --- | --- |
| **Age, y** |  |
| Median | 42 |
| Range | 18-84 |
| **Sex** |  |
| Male | 50 (59) |
| Female | 35 (41) |
| **Race** |  |
| African American | 3 (3.5) |
| Asian | 7 (8.2) |
| Hispanic | 30 (35) |
| White | 45 (53) |
| **Sample collection** |  |
| Pre-treatment only | 42 |
| Pre-treatment & Post-treatment | 28 |
| Post-treatment only | 15 |
| **WBC count, X 10<sup>9</sup>/L** |  |
| Median | 4.2 |
| Range | 0.1-107.3 |
| **Salvage-treatment phase** |  |
| zero | 4 (4.7) |
| First | 34 (40) |
| Second | 25 (29) |
| Third | 9 (11) |
| Fourth | 8 (9.4) |
| Fifth | 5 (5.9) |
| **BM blast percentage, n (%)** |  |
| Median (range) | 66 (10-97) |
| >5% to ≤50% | 28 (33) |
| >50% | 53 (62) |
| Unknown | 4 (4.7) |

Overall, 51 of 85 patients (60%) achieved CR/CRi with InO treatment. We investigated whether genomic subtypes were associated with response to InO. Genetic subtypes were determined by analysis of gene rearrangements, DNA copy number alterations, somatic single nucleotide variation (SNV) or insertion/deletion (indel) mutations and gene expression data (Supplementary Table 1). The most common (>10%) subtypes were *BCR::ABL1*-like (29%), *BCR::ABL1*-positive (14%), low hypodiploid (13%), and *KMT2A*-rearranged (*KMT2A*-R, 11%), consistent with the known distribution of genomic subtypes observed in adult B-ALL.^16^ Less frequent B-ALL subtypes (<5% each) were grouped together as “other subtypes” (n=23) for outcome analysis, and comprised *TCF3*::*PBX1*, *DUX4*-rearranged, *BCL2*/*MYC*, *CDX2*/*UBTF*, hyperdiploid, *MEF2D-*rearranged, *ZNF384*-rearranged, *ETV6*::*RUNX1*, *ZEB2*/*CEBP*, *PAX5* P80R, and unclassified (B-other) cases. The “other subtypes” were predominantly standard-to-intermediate risk based on molecular classification of risk assessment, while *BCR::ABL1*-like, low hypodiploid and *KMT2A*-R were considered high risk.^16^ As expected, the CR/CRi rates were generally lower in the *BCR::ABL1*-positive, *BCR::ABL1*-like, low hypodiploid and *KMT2A*-R subtypes than in “other subtypes” (Supplementary Table 2). The event-free survival (EFS) and overall survival (OS) rates varied among subtypes (Supplementary Figure 1A-B). Compared to “other subtypes” group, EFS was shorter in *BCR::ABL1*-positive (*P*=0.02) and low hypodiploid (*P*=0.006), but the differences were not significant in *BCR::ABL1*-like (*P*=0.5) or *KMT2A*-R (*P*=0.1).

As *KMT2A*-R ALL may undergo lineage switch following CD19-directed therapy^17–19^, we examined the expression of CD22, CD19 and myeloid lineage markers in *KMT2A*-R patients with post-InO samples available (n=3, 2 of which also had paired pre-InO samples). The *KMT2A*-R cases demonstrated post-InO reduced *CD22* RNA expression (47%-59% decrease) and percentage of CD22+ blasts (75%-90% decrease), with no change in the level of CD19 (supplementary Figure 1C). One sample (SJALL079501) had elevated RNA expression of myeloid lineage genes (*MPO*, *ITGAM* [CD11b], *ANPEP* [CD13], *CD33*, *CD14*) and the stem cell marker *KIT* (CD117), without detectable changes of protein expression on flow cytometry or lineage switch at post-InO relapse. This patient experienced lineage switch and relapse with acute myeloid leukemia after subsequent therapy with blinatumomab. No signs of lineage switch were observed for the other two *KMT2A*-R samples.

### Multiple mechanisms of CD22 antigen escape

No somatic *CD22* mutations were observed in pre-InO baseline samples. After InO treatment, acquired *CD22* mutations were present in 11% (3/27) of post-InO relapsed samples, but not in post-InO refractory samples (0/16). One patient with acquired *CD22* mutations (SJALL058834) was treated with InO monotherapy, and the other two (SJALL058975 and SJALL074541) with InO in combination with low-intensity chemotherapy. In all three cases, the relapse occurred after completion of InO and after subsequent HSCT. There were multiple *CD22* mutations per sample (range 3-4) with a total of 10 mutations identified (Table 2). Mutations occurred in the extracellular and cytoplasmic domains of CD22, with exon 2 that encodes the first of the 7 immunoglobin (Ig) domains as the mutational hotspot (Figure 2A). The identified mutation types included frameshift indel (n=4), nonsense SNV (n=2), missense SNV (n=3) and stoploss SNV (n=1) (Figure 2B). The sum of variant allele frequency (VAF) was proportional to the blast% (Table 2), suggesting all or nearly all cells harbored a *CD22* mutation. We identified multiple mechanisms of CD22 escape by genetic mutations as described below, including protein truncation, epitope alteration, and protein destabilization.

**Figure 2.**
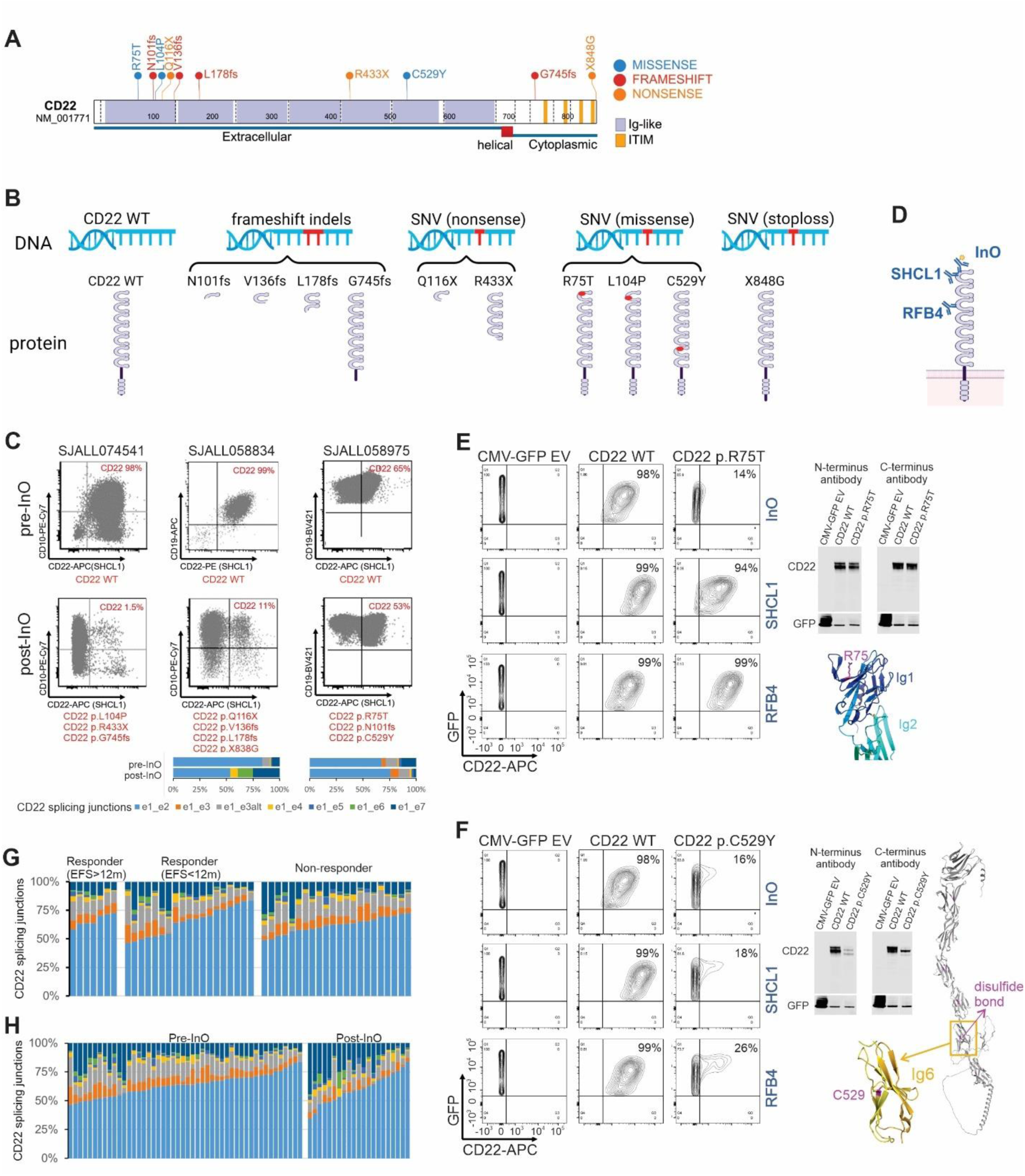
Multiple mechanisms of CD22 antigen escape. (A) Protein domain plot of all post-InO acquired CD22 mutations (n=10) identified in this cohort. (B) Wild-type (WT) and the predicted mutant CD22 protein structures. Grouped by mutation types. (C) Clinical flow plots of paired pre- and post-InO patient samples. (D) Antibodies directed to different CD22 epitopes (InO and SHCL1 that bind the first Ig domain of CD22, and RFB4 that binds the third). (E-F) Functional characterization of CD22 p.R75T and p.C529Y. CD22 protein expression and binding were performed using antibodies directed to different CD22 epitopes. (G) Stack plots depicting relative abundance of alternative splicing of CD22 exon1 in pre-InO baseline samples, comparing responders (EFS>12m and EFS<12m) and non-responders. (D) Stack plots depicting relative abundance of alternative splicing of CD22 exon1, comparing pre-vs post-InO samples.

**Table 2.**
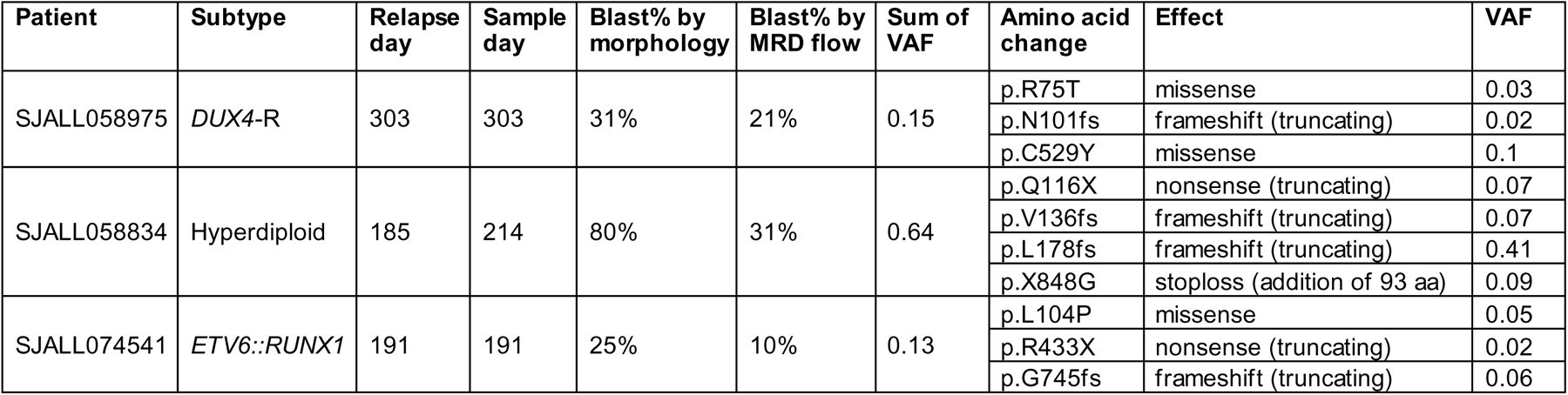
Post-InO acquired CD22 mutations.

#### Frameshift or nonsense mutations lead to CD22 protein truncation and loss

Three frameshift mutations (p.N101fs, p.V136fs, p.L178fs) and two nonsense mutations (p.Q116X, p.R433X) were predicted to generate truncated CD22 proteins lacking the transmembrane domain, and consequent a lack of membrane anchorage and loss of CD22 surface antigen (Figure 2B). Clinical flow cytometry of samples from the two patients (SJALL074541 and SJALL058834) confirmed that the post-InO samples exhibited almost complete loss of CD22 surface protein (Figure 2C).

#### Mutational alteration of the CD22-InO binding epitope without protein loss

Thepost-InO sample of SJALL058975 harbored three *CD22* mutations (CD22 p.R75T, p.N101fs and p.C529Y), but loss of CD22 expression at post-InO was not evident by clinical flow (Figure 2C). We cloned the CD22 p.R75T missense mutation and expressed it in 293T cells, and then measured CD22 protein expression and binding using antibodies directed to different CD22 epitopes (InO and SHCL1 that bind the first Ig domain of CD22, and RFB4 that binds the third Ig domain, Figure 2D). Using immunoblotting, there was no change of total CD22 protein level, but on flow cytometry, CD22 p.R75T resulted in nearly complete abrogation of InO binding (Figure 2E). The binding of CD22 by antibody SHCL1 was slightly reduced, and binding by antibody RFB4 was unchanged (Figure 2E). Protein structure modeling revealed that CD22 p.R75 was surface exposed on the first Ig domain and the energetic consequence of the p.R75T mutation was predicted to be minimal. Thus, rather than causing loss of antigen expression, CD22 p. R75T is expected to alter the InO epitope and impair InO binding. Importantly, the antibody used for clinical flow (SHCL1) still recognizes CD22 p.R75T, and thus would underestimate the degree of CD22 antigen escape to InO.

#### Mutational destabilization resulting in loss of CD22 protein

Protein expression of CD22 p.C529Y, also identified in SJALL058975, was much lower than wild type CD22, despite similar transfection rates and vector expression level (IRES-mediated expression of GFP as internal control, Figure 2F). Flow cytometry confirmed loss of binding to InO, and antibodies SHCL1 and RFB4. The CD22 protein has a “bead-on-string” structure, with 7 Ig domains as 7 beads, and each of the Ig domains is stabilized by a disulfide bond.^20^ CD22 p.C529 formed a disulfide bond in the core of the sixth Ig domain, and CD22 p.C529Y was predicted to be massively destabilizing. Using FoldX^21^, p.C529Y was predicted to destabilize this domain by 28.8 kcal/mol (wild type Ig6 was predicted to have a free energy of folding of -5.2 kcal/mol and this value for the p.C529Y mutant was predicted to be +23.6 kcal/mol), which explains the low protein level as misfolded proteins are usually degraded by the cellular quality control machinery.

#### Alternative splicing associated with downregulation of CD22 protein expression

The pre-InO sample of SJALL058975 did not have any *CD22* mutations but was partial CD22+ (65%, Figure 2C), indicating CD22 protein level were modulated by mechanisms other than genetic mutations. We performed splicing analysis as alternative splicing of exon 1 (e1) to alternative exons or junctions (e1_e3, e1_e3alt, e1_e4, e1_e5, e1_e6, e1_e7), and skipping of AUG-containing exon 2 (e2) has been reported to modulate CD22 protein expression in pediatric B-ALL.^10^ Alternative splicing of *CD22* exon 1 was observed in all patients at pre-InO baseline at various levels (16%-54%), though at the cohort level it did not appear to be a predictor of response to InO (Figure 2G). Compared to pre-InO, an increase of post-InO *CD22* exon 1 alternative splicing was observed (median 36% [range, 16%-54%] vs 41% [16%-66%], Figure 2H), however the lack of paired samples limited the interpretability of these data. Alternative spliced isoforms of exon 1 comprised 33% of *CD22* transcripts of SJALL058975 at pre-InO (Figure 2C), partly explaining reduced CD22 positivity. The post-InO sample from patient SJALL058834 had enriched alternative splicing (46% of *CD22* transcripts) in addition to multiple acquired *CD22* mutations (Figure 2C), all of which contributed to post-InO CD22 antigen escape.

### *TP53* mutations were associated with primary and acquired resistance to InO

We characterized baseline genomic alterations in genes known to be mutated in leukemia (Figure 3A). Alterations in transcriptional regulators (excluding B-lineage transcription factors) were more common in responders than non-responders (54% vs 23%, *P*=0.01). Samples from non-responders were enriched with alterations in pathways of cell cycle (77% vs 54% in responders, *P*=0.05) and Ras (35% vs 13%, *P*=0.04). Notably, *TP53* mutations were more common in non-responders than responders at pre-InO baseline (48% vs 23%, *P*=0.04, Figure 3A). *TP53* mutations were identified in 34% (24/70) of the cases with pre-InO samples. Although 9 of *TP53*-mutated cases (37.5%) attained CR/CRi, 8 relapsed within 12 months, and *TP53*-mutated cases had shorter EFS and OS comparing to cases without *TP53* mutations (Figure 3B). Among patients who were refractory or relapsed after InO, *TP53* mutant clones emerged or expanded, suggesting *TP53* mutations promoted acquired resistance to InO. Of 14 cases with *TP53* mutations in post-InO samples, two acquired *TP53* mutations post InO (one was *KMT2A*-R, two mutations, sum of VAF 0.7; one was *ZNF384*-rearranged, VAF 0.39, Figure 3C); three had *TP53* mutation VAF increased post InO (Figure 3C); and in 9 cases, mutations were hemizygous and/or maintained high VAF during disease progression (Supplementary Figure 2).

**Figure 3.**
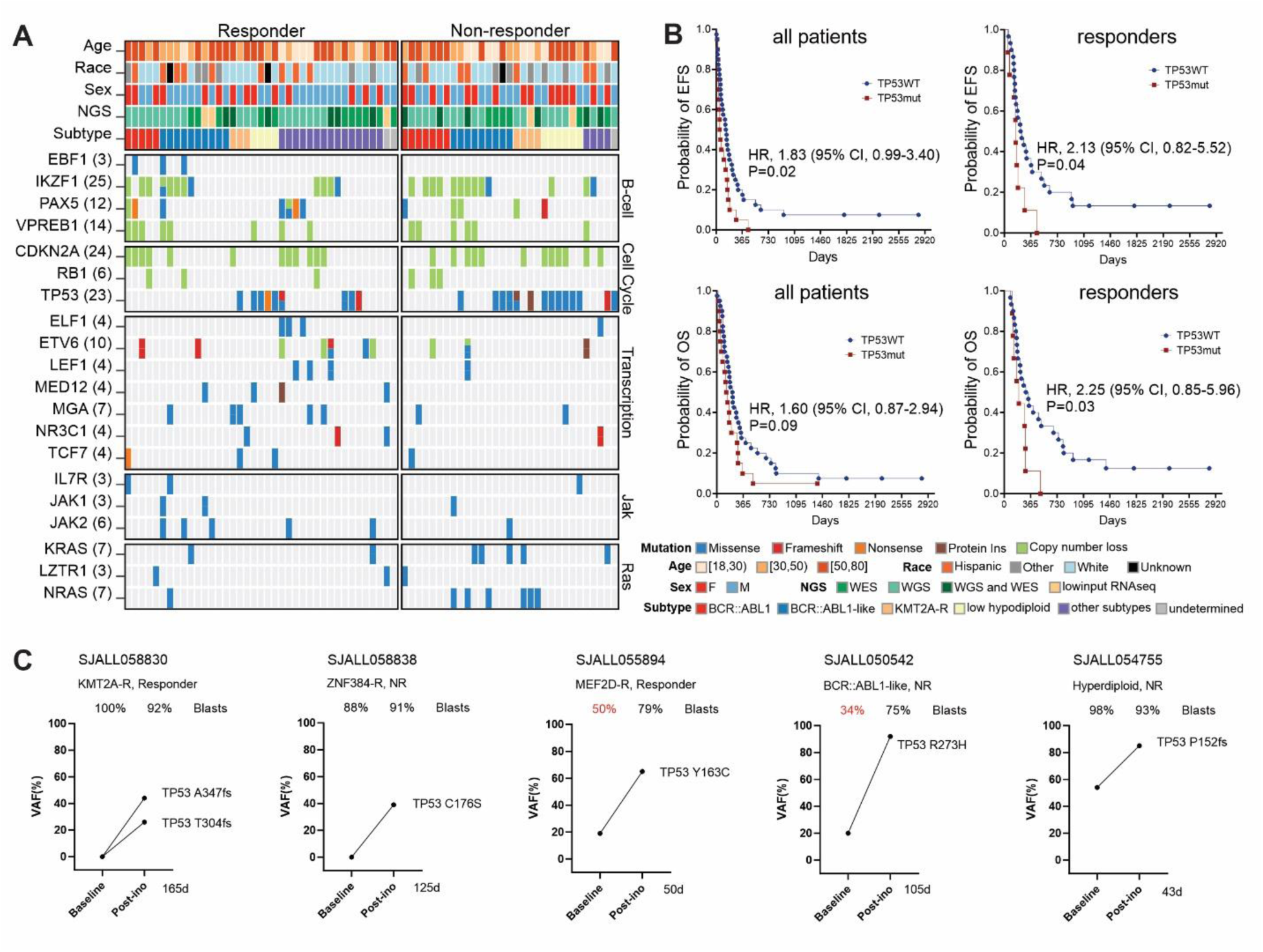
*TP53* mutations in primary and acquired resistance to InO. (A) Baseline genomic alterations in major leukemia genes, comparing responders vs non-responders. (B) EFS and OS of TP53WT vs TP53mut patients. The elapsed observation time is plotted on the curve as a circle. Symbols for both events and censored observations are plotted so that each subject was shown. Censored observations can be clearly seen as circles along the horizontal portion of the curve. Medians were estimated with the Kaplan-Meier method. *P* values were determined by log-rank test. (C) Dynamic changes in TP53 *VAF* in 5 cases with paired pre- and post-InO samples. The VAF for each mutation was shown, along with the sample blast count at the corresponding time point. The time elapsed from start of InO to treatment failure (relapse/refractory) was shown in days. EFS, Event-free survival; OS, overall survival; VAF, variant allele frequency.

### Hypermutation-driven CD22 antigen escape from InO

We examined the number and type of post-InO acquired mutations to investigate which DSBs repair pathways were leveraged to mitigate InO toxicity. Single base substitutions (SBS) were the primary mutation type in all but one patient (Figure 4A), suggesting in most cases the rapid but indel-prone DSBs repair pathways (NHEJ, a-EJ or SSA) was not primarily employed for InO-induced DNA damage; instead, HR-mediated repair, a higher fidelity mechanism, may have been utilized. Based on the density histogram of the number of acquired mutations per sample in our cohort (Figure 4A), we divided patients into post-InO low-mutators (n=21, 36-252 acquired mutations), moderate-mutators (n=3, 447-759 acquired mutations), and hyper-mutators (n=2, 1737-2433 acquired mutations). Patients who were refractory to InO and those who relapsed during InO therapy were all low-mutators; in contrast, patients who relapsed after completion of InO were low-, moderate-, or hyper-mutators (Figure 4A). Notably, the two hyper-mutators (SJALL058834 and SJALL074541) and one moderate-mutator (SJALL058975) acquired multiple *CD22* mutations post-InO (Figure 4A and 4B), suggesting accelerated acquisition of mutations was one driving force behind CD22 antigen escape to InO.

**Figure 4.**
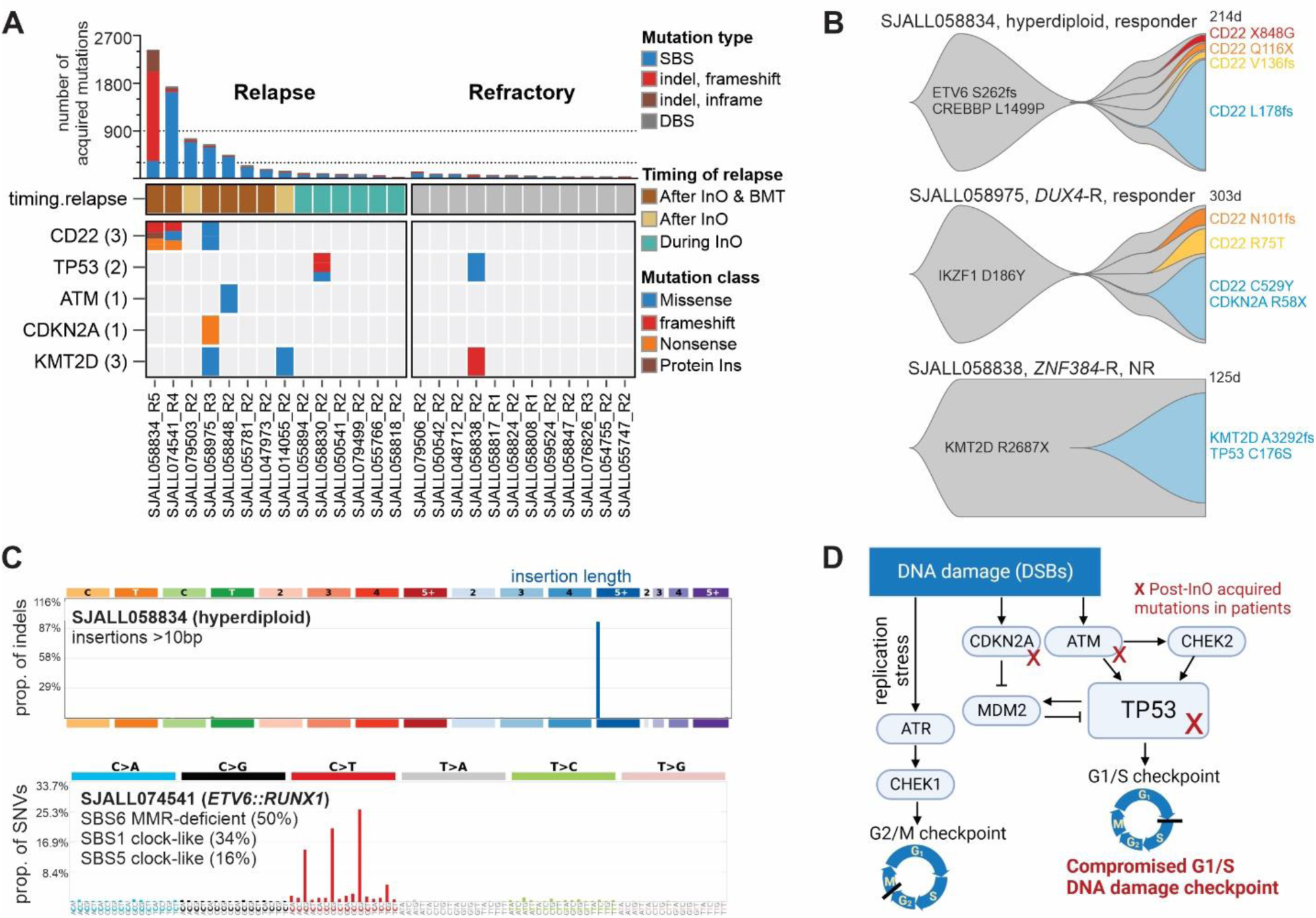
Post-InO acquired determinants of resistance. (A) The number (top) and oncoprint (bottom) of post-InO acquired mutations. The dotted lines (at 300 and 900, number of post-InO acquired mutations) were used as cutoffs to divide patients into post-InO low-mutators, moderate-mutators and hyper-mutators. (B) Fishplots showing acquisition of mutations in CD22 and KMT2D. (C) The mutational signatures of post-InO acquired mutations in hyper-mutators SJALL058834 (>10bp insertions) and SJALL074541 (SBS6, MMR-deficient). (D) A simplified schematic of compromised G1/S DNA damage checkpoint by acquired mutations in InO-treated patients. Created with BioRender.com.

Mutational signature analysis revealed that the hypermutation was generated either by a-EJ repair in response to InO-induced DSBs, or by tumor-intrinsic mismatch repair (MMR) deficiency. The primary mutation type of post-InO acquired mutations in hyper-mutator SJALL058834 were indels (86%, Figure 4A), most commonly insertions of >10 nucleotides (74%, Figure 4C). These features were not present in its pre-InO sample (7% mutations were indels, 1.7% were insertions >10 nucleotides), indicating this post-InO hypermutation signature was treatment induced. Of the four DSBs repair pathways, a-EJ was known to create long insertions,^11^ suggesting SJALL058834 tumor cells might have resisted InO through a-EJ DNA repair. In contrast, hyper-mutator SJALL074541 acquired SBS6 signature mutations attributable to defective MMR (Figure 4C). Its pre-InO sample harbored a loss of function mutation in *MLH1* (MLH1 p.L155fs) and was therefore tumor-intrinsic MMR deficient, which was recognized as the cause of hypermutation and corresponding mutational signature.

### Resistance to InO by compromised G1/S checkpoint or mitigation of replication stress

We observed acquired loss-of-function mutations post-InO in genes that regulate the G1/S DNA damage checkpoint, in addition to the previously mentioned *TP53* mutations (Figure 4D). SJALL058848 acquired missense mutation ATM p.V2424G, a known loss-of-function pathogenic variant that impairs ATM kinase activity.^22^ SJALL058975 acquired the nonsense p.R58X mutation in the CDKN2A tumor suppressor upstream of TP53. As the G1/S checkpoint is often compromised during tumorigenesis,^23^ our observations suggested that acquired mutation-induced defects in G1/S DNA damage checkpoint contribute to InO resistance. Importantly, as tumor cells with compromised G1/S checkpoint depend on the G2/M checkpoint,^23^ this vulnerability may be exploited through the use of selective molecularly targeted agents (e.g., G2/M checkpoint inhibitors) in combination with InO.

Another frequently post-InO mutated gene was *KMT2D* (*MLL4*, n=3, Figure 4A). SJALL058838 (*ZNF384*-rearranged) had a pre-existing loss-of-function KMT2D mutation (p.R2678X) and acquired another mutation (KMT2D p.A3292fs) post-InO, resulting in disruption of both *KMT2D* alleles (Figure 4B). Loss of KMT2D inhibits recruitment of the MRE11 nuclease to stalled replication forks, resulting in replication fork protection and cell survival by mitigation of replication stress; this is a known resistance mechanism to DNA damaging agents such as PARP inhibitor.^24^

### Genome wide CRISPR screening identified InO resistance and sensitization targets

We undertook genome wide CRISPR/Cas9 screening of the NALM-6 (*DUX4-*rearranged) cell line to understand genomic determinants of response and resistance to InO. Using false discovery rate (FDR) <0.1 at passage 5 (P5) and/or passage 10 (P10), we identified 274 genes whose loss led to InO sensitization (negatively selected) and 102 genes whose loss led to InO resistance (positively selected) (Supplemental Figure 3A, Supplementary Table 3). Gene set enrichment analysis (GSEA) identified multiple pathways relevant to InO sensitization, including DNA damage and DNA repair pathways (Supplemental Figure 3B). In addition, *ABCC1*, encoding an ABC transporter protein, was among the top sensitizing targets (Supplemental Figure 3A), suggesting ABCC1 mediated drug efflux was an important mechanism for InO clearance. Interestingly, *DNTT* (encoding terminal deoxynucleotidyl transferase, TdT) came out as the top resistance gene (Supplemental Figure 3A, 3C). The canonical function of TdT is to generate lymphocyte antigen receptor diversity by adding nontemplated nucleotides to DNA 3′ terminal ends during V(D)J recombination.^25^ Recent studies demonstrated that TdT was the key factor driving the DSBs repair outcome, and *DNTT* knockout in Jurkat cells shifted insertion-dominant repair to deletion-dominant repair.^26^ Another study reported that TdT-negative T cell acute lymphoblastic leukemia/lymphoma (T-ALL/LBL) patients had poor therapeutic responses; TdT loss decreased apoptosis, induced the accumulation of chromosomal abnormalities and tolerance to abnormal karyotypes.^27^ *DNTT* loss might modulate the DSBs repair response to InO and result in increased tolerance to InO-induced DNA damage, leading to InO resistance. We observed decreased *DNTT* RNA expression in patient samples at post-InO (*P*=0.03, mean of different -1.19 log2[counts per million], 95% CI [-2.23, -0.11], Supplemental Figure 3D); no post-InO acquired *DNTT* mutations were identified in this dataset. TdT is commonly utilized in clinical flow and can be prospectively evaluated as a novel marker of InO response. *CD22* arose as the resistance gene as expected. Moreover, loss of *CHEK2*, the crucial G1/S checkpoint gene, led to InO resistance (Supplemental Figure 3A and 3C). This supported the observation and hypothesis from patient data (Figure 4D) that tumor cells resisted InO by compromise of the G1/S DNA damage checkpoint.

## DISCUSSION

CD22 is broadly and uniquely expressed by B-lineage cells and therefore an attractive candidate for targeted therapy in B-ALL. Our study shed light on mechanisms of CD22 antigen escape. Similar to CD19,^8,9^ CD22 escape could occur by acquired truncating mutations; moreover, this escape could occur through the alternation of just one amino acid. *CD22* missense mutations could be massively destabilizing and result in CD22 protein loss (e.g., CD22 p.C529Y), or be epitope-specific without causing CD22 protein loss or affecting binding to other antibodies (e.g., CD22 p.R75T). The latter type of mutation poses a challenge on how to accurately monitor for CD22 antigen escape. Ideally, not only “antigen loss”, but also “antigen-antibody binding” needs to be evaluated. For clinical flow, multiple CD22 antibodies directed to different epitopes, including an antibody with the same/similar epitope as InO, could be utilized to monitor for CD22 escape. In our cohort, acquired *CD22* mutations were only observed in cases relapsed to InO (rather than refractory), and *CD22* mutations occurred at relatively low prevalence (11% of the relapsed cases), suggesting the alternative mechanisms drive resistance apart from direct mutational perturbation of the target antigen. First, hypermutation by tumor-intrinsic MMR deficiency or InO-induced rapid low-fidelity DNA damage repair drove accelerated acquisition of mutations, leading to selection for clones with survival advantages (including *CD22* mutations). Second, mutations of crucial genes or pathways could abrogate DNA damage induced apoptosis, resulting in calicheamicin resistance. Patients with *TP53* mutations were reported to achieve high CR/CRi rates in response to InO in two cohorts with a small number of *TP53*mut cases.^28,29^ In our cohort, the response rate in *TP53*mut patients was lower (9/24 [37.5%]). Notably, new *TP53* mutant clones emerged at post-InO treatment, suggesting that *TP53* mutations mediate both primary and acquired resistance to InO. Of the DNA damage checkpoints, the G1/S checkpoint is unique in depending primarily on the function of p53 .^13,14^ In addition to *TP53* mutations, post-InO acquired mutations in other G1/S DNA damage checkpoint genes (*ATM* and *CDKN2A*) were observed in tumor samples, and loss of *CHEK2* resulted in InO resistance in NALM-6 cell line, suggesting tumor cells evaded InO-induced apoptosis by compromising the G1/S checkpoint. These observations, though small in numbers, justified further investigations in DNA damage checkpoints for determinants of response to InO. We observed a striking association of *DNTT* loss and InO resistance in the CRISPR screen, advocating for prospective evaluation of *DNTT* as a new biomarker of InO resistance.

One limitation worth considering is that some patients received InO in combination with low-intensity chemotherapy, and therefore some relapses may not have been mediated by InO-based mechanisms of resistance (rather the patients may have become resistance to the backbone chemotherapy). Nevertheless, we observed two of these patients still acquired *CD22* mutations post-treatment, suggesting InO-mediated resistance during disease recurrence in patients treated with combinational therapy.

In summary, we demonstrated the novel mechanisms of mutation-induced CD22 escape including protein loss and epitope alteration, identified the genomic features driving *CD22* mutagenesis, and described the DNA damage response relevant to calicheamicin resistance. These observations contribute to our understanding of escape strategies within and beyond antigen loss to CD22-targeted therapy, which may lead to improved therapeutic approaches in the future.

## METHODS

### Patients and clinical specimens

We retrospectively studied 85 adult B-ALL patients treated at MD Anderson Cancer Center. The selection criteria were patients with pre-InO and/or post-InO tumor samples (banked cells/DNA/RNA from bone marrow or peripheral blood-derived tumor cells) available. The study was approved by the institutional review boards of MD Anderson Cancer Center and St Jude Children’s Research Hospital, with informed consent.

### Transcriptome sequencing

Total stranded transcriptome sequencing (RNA-seq; 100bp paired-end reads) was performed using the TruSeq Stranded Total RNA library preparation kit and sequencing performed using HiSeq 4000 or NovaSeq 6000 platforms (Illumina) to generate a minimum of 100 million reads per sample library with a target of greater than 90% mapped reads. The low input RNA library preparation kit (NuGen Ovation V2) was used for samples with limiting material (2-100 ng). The adapters in sequencing reads were trimmed with Trim Galore (v0.4.4; https://www.bioinformatics.babraham.ac.uk/projects/trim_galore/, -q 20 –phred 33 –paired). The trimmed sequencing reads were mapped with STAR (v2.7.9a)^30^ to human genome GRCh38. The expected gene counts calculated using RSEM^31^ for each sample were compiled to one gene count matrix. Only genes annotated as level 1 or 2 by GENCODE (v31) were kept in the downstream analysis.

### Whole genome and exome sequencing

Whole exome sequencing (WES) of genomic DNA was performed using the TruSeq DNA Exome library preparation kit (Illumina). Sequencing (100bp paired-end reads) was performed using the NovaSeq 6000 platform (Illumina) to an minimum haploid coverage of 100x. Whole genome sequencing (WGS) libraries were prepared using the HyperPrep library preparation kit (Roche) and sequenced using the NovaSeq 6000 System (Illumina) to a target depth of 800 million paired-end 150bp reads per sample for 30x average haploid coverage. The paired end sequencing WGS and WES reads were mapped with BWA-MEM^32^ to human genome GRCh38. The alignment quality was assessed using Qualimap. Genomic data is publicly available and has been deposited in the European Genome Phenome Archive, Accession EGAS50000000067.

### Genetic subtyping

Subtypes were determined by integrating gene expression, rearrangement, copy number and SNV/indel as previously described.^33,34^. Samples without RNA-seq were usually assigned to the B-other subtype; however, in cases without RNA-seq but with WGS, subtype-defining rearrangements if detected by WGS were used to assign subtypes. Cytogenetic and FISH data collected on the clinical trials was compared against the results of genomic analyses, and discrepancies were resolved by in depth review.

### Mutation analysis

Variants were called by Mutect2 (v4.1.2.0)^35^ using the tumor-only calling approach. Multiple filtering steps were applied to exclude potential calling artifacts. The variants passing the filtering steps fulfilled the following criteria: coverage depth of the variant > 10, variant allele frequency > 0.01, alternative allele count > = 4, allele population frequency in public databases < 0.01 (gnomadAD, 1000 genomes, ExAC and Exome Sequencing Projects), mappability > 0.7, not co-localized with repeat elements and GC percentage between 0.4 and 0.6. The variant annotation was performed using Annovar.^36^ Oncoprint was generated using ProteinPaint.^37^ Post-InO acquired mutations were identified by using pre-InO sample to filter its paired post-InO samples. Two-dimensional MAF plots were used to visualize the relationship between pre- and post-InO samples. Clonal evolution was visualized using fishplot.^38^ Post-InO acquired mutations were used to analyze post-InO mutational signatures. Mutational signatures were profiled by fitting SNV and indel counts per 96 tri-nucleotide contexts to the COSMIC signatures version 3.2. Signatures with <5% overall contribution were excluded from the summary.

### Copy number analysis using whole genome sequencing

Chromosome level copy numbers were estimated using coverage analysis of WGS. Control-FREEC (Control-Free Copy number caller)^39^ software was used to estimate genome-wide copy-numbers using the mode without control sample. Read counts were corrected by GC content and mappability. Window size was automatically adjusted using coefficientOfVariation value of 0.08. Boundaries of focal copy number alterations (CAN) were modified if they overlapped with a structure variant call from Delly (v0.7.7).^40^ Recurrent CNAs were analyzed using GISTIC2. ^41^

### *CD22* mutations modeling and characterization

The protein structural model of full-length CD22 was obtained from AlphaFold2^42^ and mutations were introduced using foldX^21^ in order to assess effects on protein stability. CMV-*CD22*-IRES-GFP vector was purchased from GeneCopoeia, Inc. *CD22* SNV/indel mutations were generated using Q5 Site-Directed Mutagenesis Kit (New England Biolabs) and the full mutation sequences were validated by Sanger sequencing. CMV-IRES-GFP empty vector, *CD22* WT vector, and *CD22* mutant vectors were transfected into HEK-293T cells using FuGENE HD (Promega). Transfected HEK-293T cells were dissociated using TrypLE Express Enzyme (Gibco, Thermo Fisher Scientific) before staining. CD22 binding experiments were performed using antibodies directed to different CD22 epitopes (supplementary Table 4). For flow cytometry, cells were stained with InO complexed with FabuLight AF647 Goat Anti-Human IgG, Fcγ fragment specific (Jackson Immunoresearch), CD22 clone SHCL1 (BD Biosciences), or CD22 clone RFB4 (Thermo Fisher Scientific). The flow gating strategy was summarized in supplementary Figure 4. Western blot was performed using antibodies specific to the N-terminus (clone E7L6Z, Cell signaling) or C-terminus (polyclonal, BosterBio) of CD22 protein and imaged using Odyssey CLx Imager (LI-COR Biosciences).

### *CD22* exon junction analysis

*CD22* junction spanning reads were obtained from the STAR “*_SJ.out.tab” result files based on its chromosome location (chr19: 35329187 – 35347361), and the exon position was annotated using *CD22* isoform NM_001771.4 by BEDTools.^43^ The *CD22* Δex2* isoforms that skipped exon2 (e2) and variably connected exon1 (e1) to several possible downstream exons (exon3 [e1_e3], exon3-alt [e1_e3alt], exon4 [e1_e4], exon5 [e1_e5], exon6 [e1_e6], exon7 [e1_e7]) were previously reported.^10^ Each entry was normalized by dividing by the total number of junction-spanning reads as previously described.^10^ Samples with low *CD22* expression (< 20 reads) were excluded from the analysis as the exon junction usage could not be accurately estimated due to low number of reads.

### Statistical analysis

Differencesbetween groupswere tested using two-sided Fisher’s exact test. Median PFS and OS were estimated using the Kaplan-Meier method, with the *P* values, hazard ratio (HR) and corresponding 95% confidence intervals (CIs) calculated using log-rank method. GraphPad Prism 8.3.0 was used for statistical analysis.

### Genome-wide CRISPR/Cas9 screen

Whole genome CRISPR/Cas9 screen was performed as previously described.^44^ Briefly, human CRISPR Knockout Pooled Library (Brunello)^45–47^ (Addgene #73179) lentivirus was obtained from the Center for Advanced Genome Engineering at St. Jude. NALM-6 cells were infected with the genome-wide sgRNA library at 20%-40% transduction efficiency to ensure no more than one sgRNA per cell, and infections were performed in three biological replicates per cell line and achieved a target representation of ∼500 cells per sgRNA. At 48 hours post transduction, transduced cells were selected by 0.5µg/ml puromycin for 7 days. After 7 days, from each replicate, at least 40×10^6^ cells were harvested for the baseline counts (Day 0 control), and remaining cells were divided into two equal groups. One group was treated with 1 ng/ml InO (half maximal inhibitory concentration, IC50) while the other group served as no-treatment control. Cells were passaged every 2-3 days with complete media change. Cells from the baseline and the endpoints (Passage 5 and 10) were subjected to genomic DNA extraction using QIAamp Blood Maxi Kit (Qiagen). Genomic DNA from each passage were amplified as per Doench et al.^45^ and sequenced. CRISPR KO screens were analyzed using MAGeCK (0.5.9.5).^48^ Pathway enrichment analysis was performed using Pre-ranked Gene Set Enrichment Analysis (GSEA) against ontology gene sets (C5 version c5.all.v2022.1.Hs.symbols.gmt) using L2FC of entire genes in the CRISPR library.

## Data Availability

All data produced in the present work are contained in the manuscript or available online at the European Genome Phenome Archive (Accession EGAS50000000067).

## ACKNOWLEDGEMENTS

We thank the staff of the Hartwell Center for Bioinformatics and Biotechnology, the Flow Cytometry and Cell Sorting Shared Resource, Center for Applied Bioinformatics, and the Center for Advanced Genome Engineering at St. Jude Children’s Research Hospital. The authors also thank Shane Marshall and William Greene at St. Jude Children’s Research Hospital for providing inotuzumab reagent for experiment, and Jairo Matthews at MD Anderson Cancer Center for shipping the patient samples. This study was supported in part by Pfizer, the National Cancer Institute of the National Institutes of Health under Award Number R35 CA197695 (C.G.M.), Cancer Center Support Grants (CCSGs) P30 CA021765, and ALSAC. C.G.M. is the William E. Evans Endowed Chair at St. Jude Children’s Research Hospital.

## AUTHOR CONTRIBUTIONS

C.G.M., K.G.R., M.K. and E.J.J. conceived and designed the study; N.J.S., E.J.J., M.K., H.M.K., W.M., N.J., B.T., J.K., G.G.M., H.Y., R.S.G. and L.F.N. provided patient samples and clinical data; Y.Z. and P.S.G. performed experiments; Y.Z., C.Q., T.C.C., P.S.G., A.H.P., W.Z., Y.F., R.W.K. and K.G.R. analyzed data; Y.Z., N.J.S. and C.G.M. prepared the manuscript; all authors reviewed and approved the manuscript.

