## Supplementary Figures for "Genomic determinants of response and resistance to inotuzumab ozogamicin in B-cell ALL"

##### Supplementary Figure legends

**Supplementary Figure 1. Response to inotuzumab by genetic subtypes.** (A-B) Event-free survival (EFS) and overall survival (OS) by genetic subtypes. Medians were estimated with the Kaplan-Meier method and P values were determined by log-rank test. The elapsed observation time was plotted on the curve as a tick. (C) Gene expression levels comparing pre- and post-InO in *KMT2A*-R patients.

**Supplementary Figure 2. *TP53* mutations were hemizygous and/or maintained high VAF during disease progression.** The VAF for each mutation is shown, along with the sample blast count at the corresponding time point. The time elapsed from start of InO to treatment failure (relapse/refractory) is shown in days.

**Supplementary Figure 3. Genome wide CRISPR screening with InO in NALM-6 cell line (passage 10).** (A) Volcano plots showing InO sensitization (negatively selected, in blue) and resistance (positively selected, in red) genes by CRISPR screen. (B) GSEA negatively enriched pathways by CRISRP screen. The color and size of the dots represent normalized enrichment score (NES) and −log10 false discovery rate (FDR) q-value of the negatively enriched pathways respectively (magma). Top 20 pathways and their respective geneset members significantly selected by CRISPR screen are denoted by bubble plot. Viridis colors and size of the dots represent running enrichment score and −log10 of CRISPR p-value respectively. (C) Log2FC values of sgRNAs targeting the top 10 InO resistance genes by CRISPR screen. Each gray line represents an individual sgRNA and the central dotted line represents median log2FC of the sgRNAs (around 0). Each red line represents a sgRNA of the target gene. Multiple sgRNAs targeting each of these top 10 genes (including CD22) were significantly enriched by inotuzumab, showing on-target gene knockout by CRISPR/Cas9. (D) *DNTT* RNA expression in paired patient samples comparing pre- vs post-InO. The plot represents mean ± standard deviation in each group. *P*=0.03 by two tailed t-test. FC, fold change; sgRNAs, single-guide RNAs; CPM, counts per million; GSEA, Gene set enrichment analysis identified.

**Supplementary Figure 4. Gating strategy for transfected cells (GFP-positive).** Cells were first visualized using FSC-A/SSC-A, followed by gating on DAPI negative population to include live cells only. Cells were gated by FSC-A/SSC-W to exclude doublets. The live singlet cells were gated for GFP positive as the transfected population, which was then evaluated for expression of the protein of interest.

#### Supplementary Figure 1

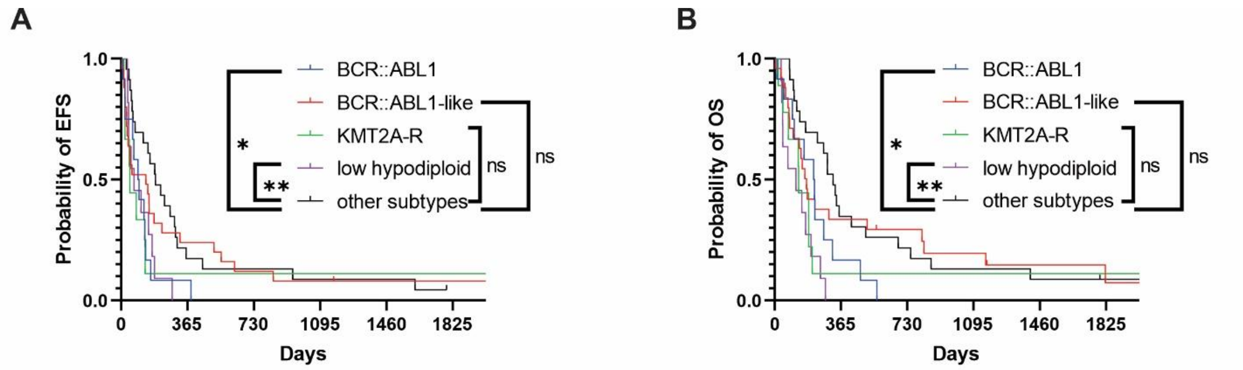


| EFS | P value | HR (95% CI) |
|---|---|---|
| BCR::ABL1/other subtypes | 0.02 | 2.22 (0.95, 5.16) |
| low hypodiploid/other subtypes | 0.006 | 2.56 (1.02, 6.45) |
| BCR::ABL1-like/other subtypes | 0.5 | 1.21 (0.67, 2.18) |
| KMT2A-R/other subtypes | 0.1 | 1.82 (0.70, 4.74) |

| OS | P value | HR (95% CI) |
|---|---|---|
| BCR::ABL1/other subtypes | 0.03 | 2.09 (0.91, 4.78) |
| low hypodiploid/other subtypes | <0.0001 | 3.59 (1.27, 10.16) |
| BCR::ABL1-like/other subtypes | 0.7 | 1.15 (0.62, 2.10) |
| KMT2A-R/other subtypes | 0.1 | 1.92 (0.72, 5.11) |

##### C. *KMT2A-R* patients

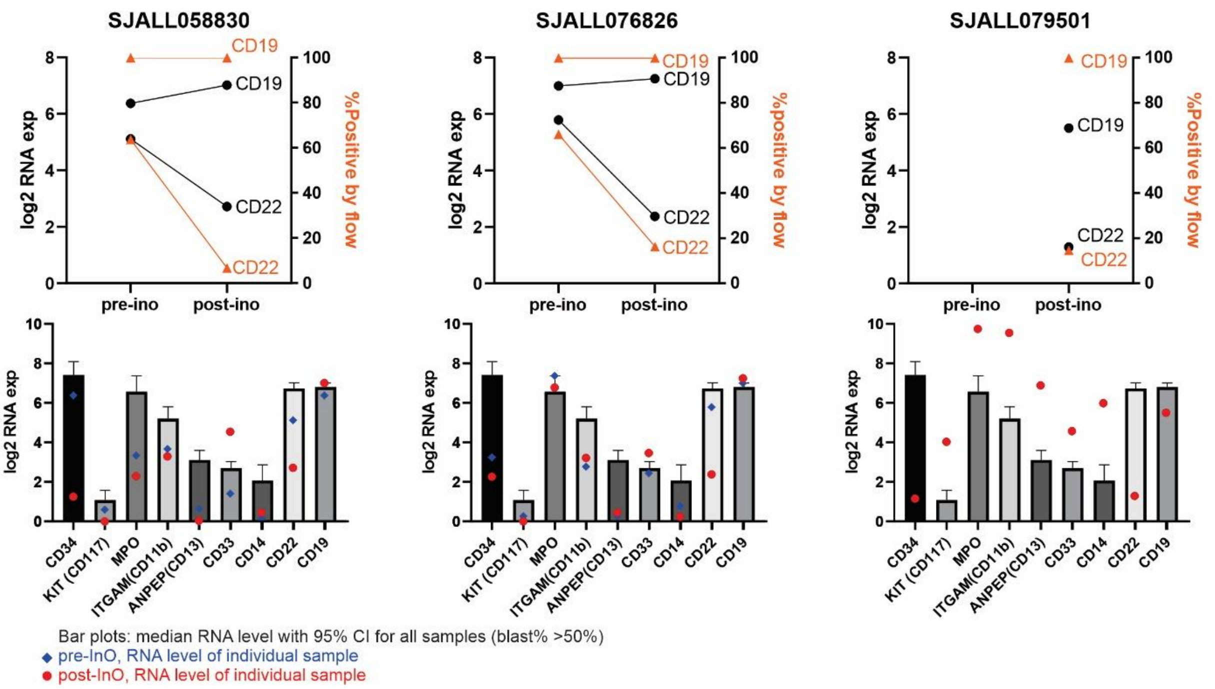

#### Supplementary Figure 2

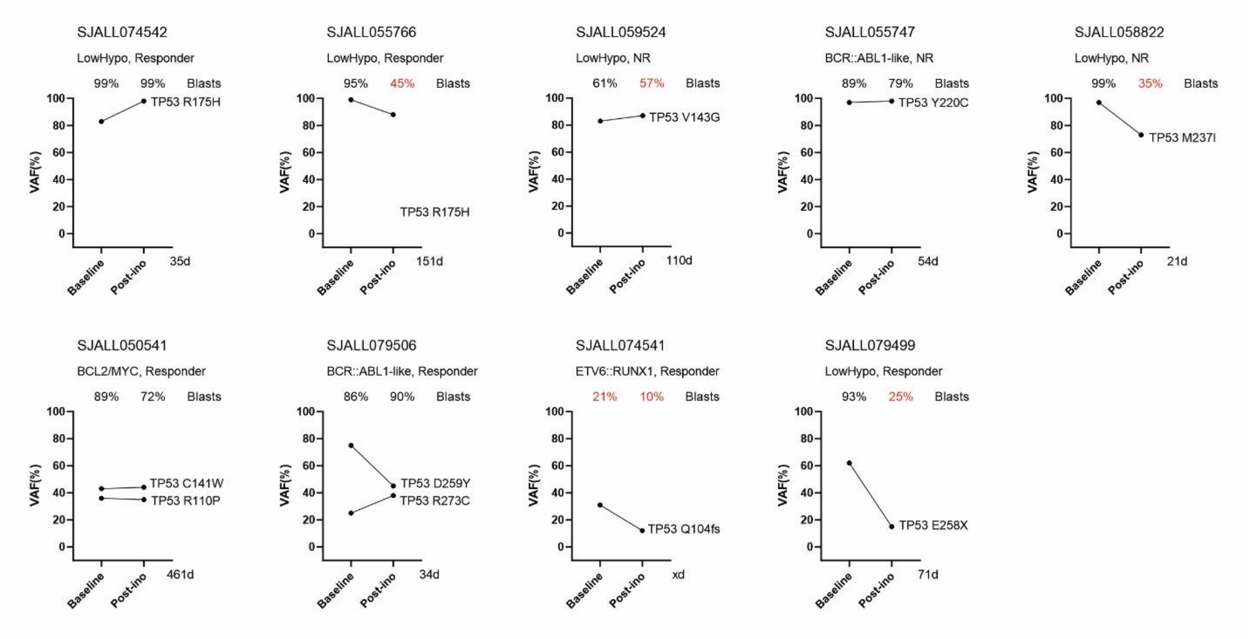

### Supplementary Figure 3

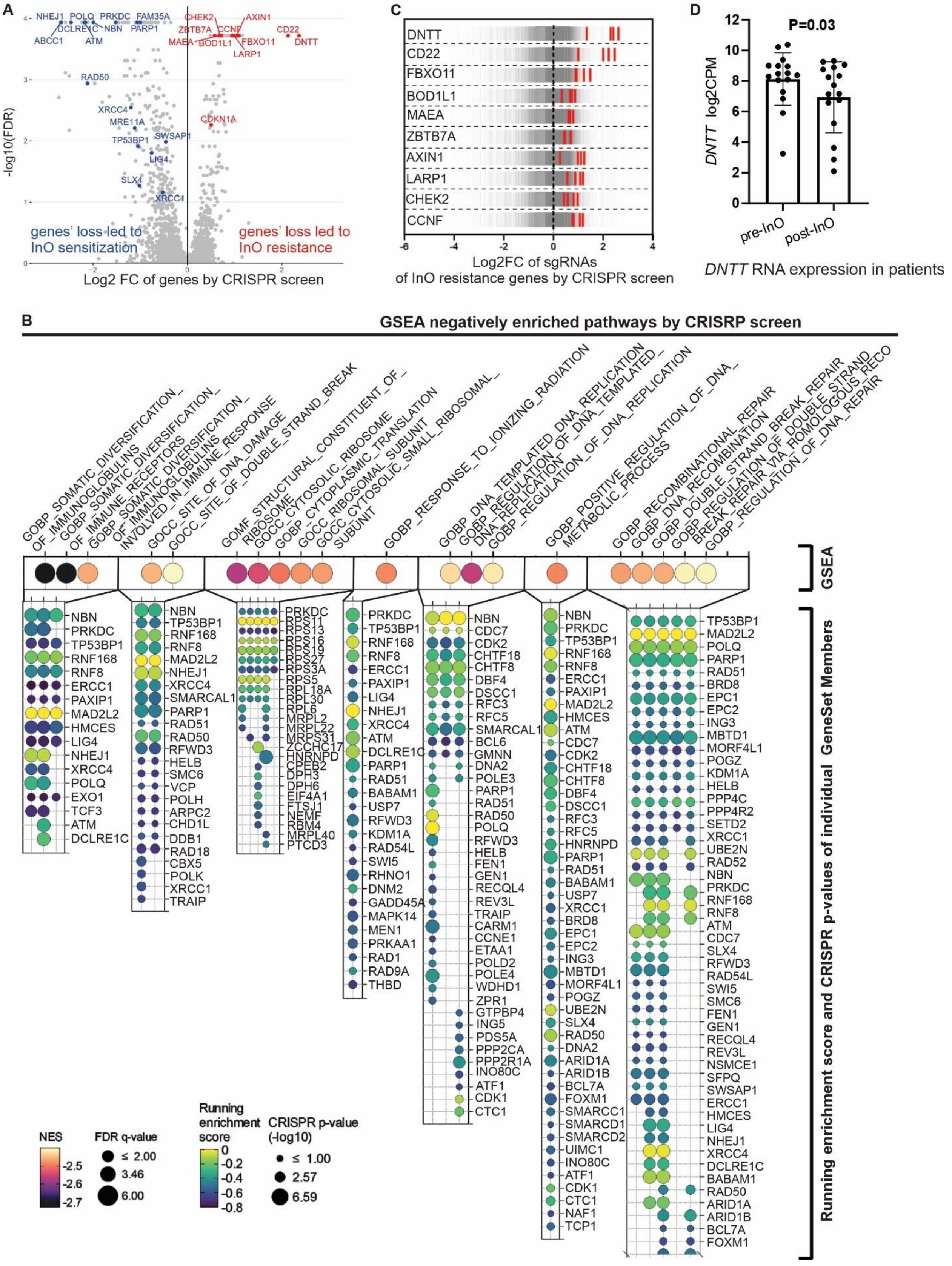

#### Supplementary Figure 4

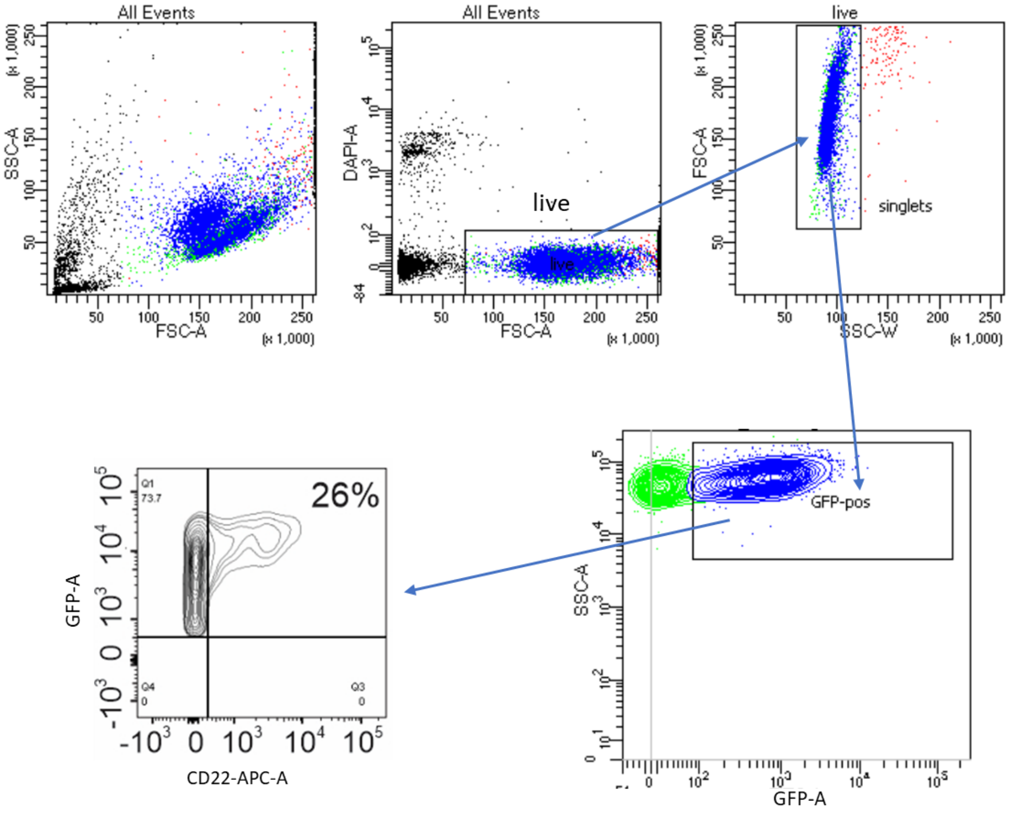
